# MedSegBench: A Comprehensive Benchmark for Medical Image Segmentation in Diverse Data Modalities

**DOI:** 10.1101/2024.08.26.24312619

**Authors:** Musa Aydin, Zeki Kuş

## Abstract

MedSegBench is a comprehensive benchmark designed to evaluate deep learning models for medical image segmentation across a wide range of modalities.. This benchmark includes 35 datasets with over 60,000 images, covering modalities such as ultrasound, MRI, and X-ray. It addresses challenges in medical imaging, such as variability in image quality and dataset imbalances, by providing standardized datasets with train/validation/test splits. The benchmark supports binary and multi-class segmentation tasks with up to 19 classes. Evaluations are conducted using the U-Net architecture with various encoder/decoder networks, including ResNets, EfficientNet, and DenseNet, to evaluate model performance. MedSegBench serves as a valuable resource for developing robust and flexible segmentation algorithms. It allows for fair comparisons across different models and promotes the development of universal models for medical tasks. The datasets and source code are publicly available, encouraging further research and development in medical image analysis.

## Background & Summary

Deep learning has become essential in medical image analysis and segmentation, offering powerful methods to help doctors and researchers better understand and diagnose diseases^1^. Deep learning can identify patterns and details in medical images that might be difficult for human eyes to detect using complex networks such as convolutional neural networks^2^. These techniques are precious for finding tumors in X-rays, classifying different cell types in whole-slide images, or segmenting different brain parts in MRI scans. However, working with biomedical datasets presents unique challenges, including variability of image quality and resolution, the need for well-annotated examples, imbalances of the datasets, and different modalities. Addressing these challenges and ensuring the effectiveness of deep learning methods in real-world medical settings requires large and diverse datasets^3^. These comprehensive collections of medical images help train the algorithms to handle different modalities and medical tasks. They also allow researchers to compare deep learning methods fairly, determine the most effective approaches for specific medical tasks, and develop universal models for different medical tasks.

There are limited benchmark studies in the literature focused on medical imaging, with most concentrating on medical image classification problems^4–8^. Gelasca et al.^4^ present a comprehensive biomedical segmentation benchmark that evaluates bioimage analysis methods. It includes six datasets with associated ground truth and validation methods, covering different scales from subcellular to tissue levels. Rebuffi et al.^5^ propose the Visual Decathlon Challenge, a benchmark that evaluates models across ten diverse visual classification domains, including datasets such as Aircraft, CIFAR-100, and ImageNet. Medical Segmentation Decathlon^6^ supports the creation and benchmarking of semantic segmentation algorithms. It includes 2633 3D images from ten different anatomical sites and modalities collected from multiple institutions and annotated by experts. Yang et al.^7^ introduce the MedMNIST Benchmark, a collection of ten pre-processed medical image datasets standardized to 28×28 pixels. It covers various medical image modalities and support multiple classification tasks. Yang et al.^8^ extend MedMNIST with MedMNIST v2, a standardized collection of biomedical image datasets. This includes 12 datasets for 2D images and 6 for 3D images, covering various data modalities, scales, and classification tasks,

This study introduces a comprehensive benchmark dataset for medical image segmentation (Figure 1). It includes 35 distinct datasets with over 60,000 images covering various data modalities such as ultrasound, dermoscopy, MRI, X-ray, OCT, and more. It provides a diverse resource for evaluating the performance of deep learning models in medical image segmentation tasks. The dataset includes a wide range of scales, from small collections with just a few dozen images to extensive datasets containing tens of thousands of samples. The segmentation tasks cover both binary and multi-class problems, with some datasets featuring up to 19 different classes. This benchmark offers several powerful advantages as a robust and versatile tool for the research community:

- **Diversity of modalities:** The benchmark includes datasets from various imaging modalities such as Ultrasound, MRI, X-Ray, OCT, Dermoscopy, Endoscopy, and various types of microscopy.
- **Task complexity:** It covers both binary segmentation tasks and multi-class segmentation tasks with up to 19 classes.
- **Dataset sizes:** There’s a wide range in the number of images per dataset, from as few as 28 to as many as 21,165.
- **Data split:** All datasets follow a standard train/validation/test split, which is crucial for the proper evaluation of machine learning models.
- **Standardization:** All datasets are standardized to enhance comparability and ease of use. Samples across all datasets have been resized to three standard resolutions - 128, 256, and 512 pixels - and stored in a uniform format.
- **Application areas:** The datasets cover various medical applications, including cancer detection, COVID-19 diagnosis, cell and nuclei segmentation, and organ segmentation.

**Figure 1.**
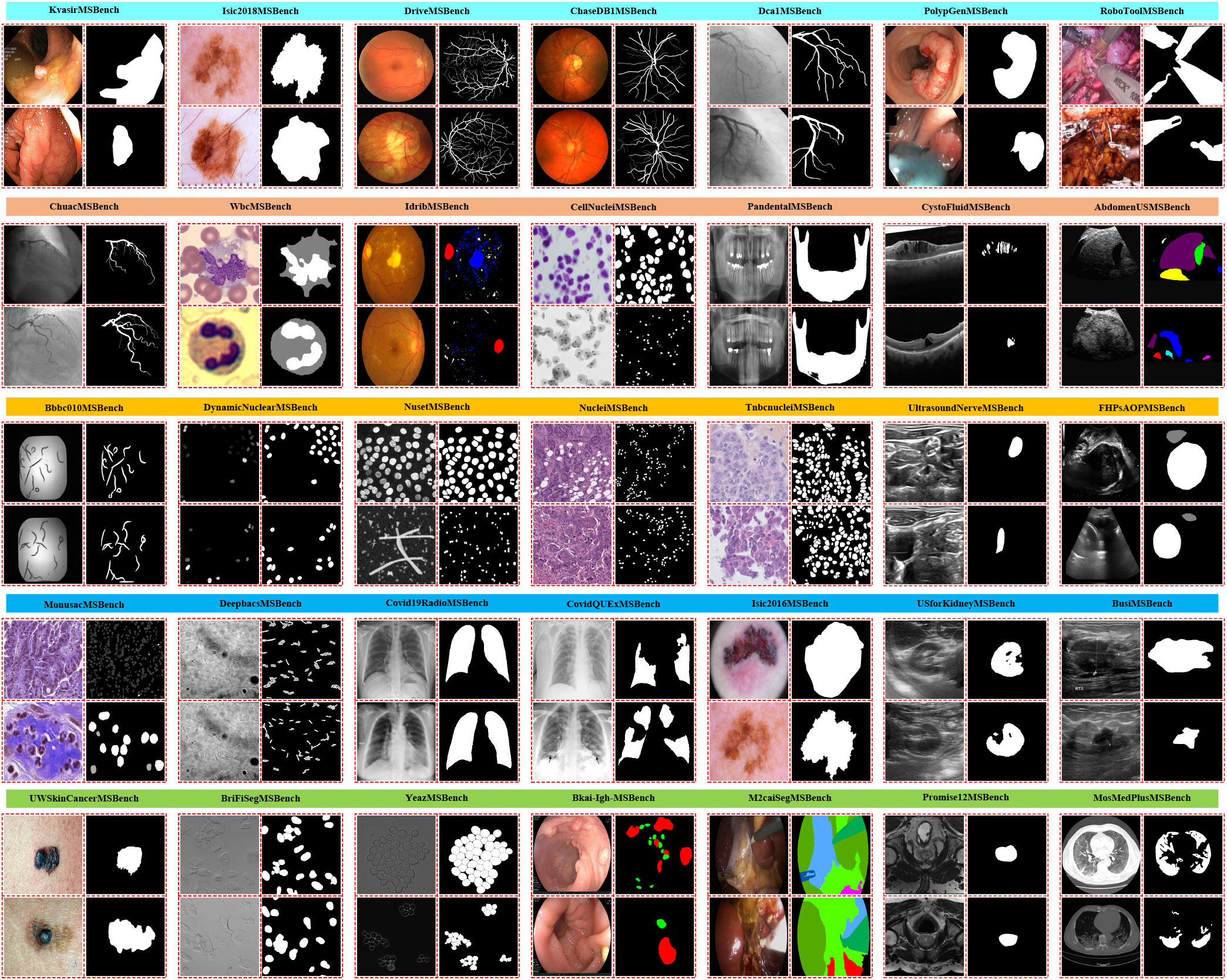
Summary of the MedSegBench.

We have evaluated each dataset on state-of-the-art segmentation model (U-Net^9^) with different encoder/decoder network types (ResNet-18, ResNet-50, Efficient-Net, MobileNet-v2, DenseNet-121, Mix Vision Transformer)^10^. Each experiment are performed 3 times and average results are reported.

This benchmark is carefully designed to assess how well deep learning models can generalize across different medical domains, perform on small and large datasets, and handle varying task complexities. By including such a wide array of medical imaging challenges, this benchmark is a powerful tool for comprehensively evaluating the robustness, flexibility, and overall efficacy of segmentation algorithms in the medical imaging field.

## Methods

### Data Preparation

The MedSegBench dataset comprises 35 distinct 2D medical image segmentation datasets, some of which are extracted from 3D slices. These datasets cover various data modalities such as Ultrasound, OCT, Chest X-ray, MR, and more. The original datasets differ in scales, segmentation tasks (binary/multi-class), classes, imaging modalities, and annotation styles. Hence, we have selected a standardized format and performed pre-processing to ensure a consistent format across all datasets.

Numerous medical image segmentation datasets are available in the literature, each presenting various challenges including variations in annotations, image sizes, and file formats. Additionally, many of these datasets lack officially shared train/test/validation splits, making it challenging to fairly compare different methods. To address these issues, we performed pre-processing steps. All image and label pairs are resized to 128×128, 256×256, and 512×512 pixels using the bicubic interpolation method. Although we used 512×512 sized images in our experiments, we have made the 128×128 and 256×256 sized versions publicly available for researchers with limited GPU memory. Also, we have applied a mapping to labels; pixels with values of 0 and 255 are mapped to 0 and 1 for binary segmentation tasks, and for multi-class segmentation tasks, pixels are mapped to integer values between 0 and (#Classes - 1). No additional augmentation or pre-processing steps are applied to the images and labels. We have followed three different scenarios based on MedMNIST v2^8^ to create train/test/validation splits: (1) Utilizing the source train/test/validation splits if published by the authors; (2) Using the source validation set as the test set and splitting the source training set into 90% training and 10% validation (9:1 ratio) if the source training and validation splits are published by the authors; (3) Randomly splitting the dataset into 70% training, 10% validation, and 20% test sets if no public train/test/validation splits are available (7:1:2 ratio). Most of these datasets are publicly published under Creative Commons Licenses, some of which are CC-BY-NC, CC-BY-SA, and CC-BY-NC-SA, permitting the redistribution of datasets. We have published datasets in MedSegBench under Creative Commons Licences, and source codes have been published under Apache License 2.0.

Table 1 presents the summary information for all MedSegBench datasets. In addition, Table 2 shows the data-modality-based overview for MedSegBench datasets. Furthermore, Table 3 provides an overview of various datasets, detailing their sub-categories and the number of samples for training, validation, and testing. In the following sections, we will describe the details of each dataset.

**Table 1.** Overview of the MedSegBench datasets, including source references, modality, task types (binary or multi-class) with number of classes, total sample sizes and train/validation/test splits.

| **Dataset Name**<sup>source</sup> | **Modality** | **Binary or Multi-class (# Classes)** | **# Images** | **# Train/Val/Test** |
| --- | --- | --- | --- | --- |
| AbdomenUSMSBench<sup>11,12</sup> | Ultrasound | Multi-class (8) | 926 | 569/64/293 |
| Bbbc010MSBench<sup>13,14</sup> | Microscopy | Binary | 100 | 70/10/20 |
| Bkai-Igh-MSBench<sup>15-17</sup> | Endoscopy | Multi-class (3) | 1,000 | 700/100/200 |
| BriFiSegMSBench<sup>18,19</sup> | Microscopy | Binary | 1,360 | 1005/115/240 |
| BusiMSBench<sup>20,21</sup> | Ultrasound | Binary | 647 | 452/64/131 |
| CellNucleiMSBench<sup>22,23</sup> | Nuclei | Binary | 670 | 469/67/134 |
| ChaseDB1MSBench<sup>24</sup> | Fundus | Binary | 28 | 19/2/7 |
| ChuacMSBench<sup>25</sup> | Fundus | Binary | 30 | 21/3/6 |
| Covid19RadioMSBench<sup>26-28</sup> | Chest X-Ray | Binary | 21,165 | 14,814/2,115/4,236 |
| CovidQUEXMSBench<sup>29,30</sup> | Chest X-Ray | Binary | 2,913 | 1,864/466/583 |
| CystoFluidMSBench<sup>31-33</sup> | OCT | Binary | 1,006 | 703/101/202 |
| Dca1MSBench<sup>34,35</sup> | Fundus | Binary | 134 | 93/13/28 |
| DeepbacsMSBench<sup>36,37</sup> | Microscopy | Binary | 34 | 17/2/15 |
| DriveMSBench<sup>38,39</sup> | Fundus | Binary | 40 | 18/2/20 |
| DynamicNuclearMSBench<sup>40,41</sup> | Nuclear Cell | Binary | 7,084 | 4,950/1,417/717 |
| FHPsAOPMSBench<sup>42,43</sup> | Ultrasound | Multi-class (3) | 4,000 | 2,800/400/800 |
| IdribMSBench<sup>44,45</sup> | Fundus | Binary | 80 | 47/6/27 |
| Isic2016MSBench<sup>46,47</sup> | Dermoscopy | Binary | 1,279 | 810/90/379 |
| Isic2018MSBench<sup>48-50</sup> | Dermoscopy | Binary | 3,694 | 2,594/100/1,000 |
| KvasirMSBench<sup>51,52</sup> | Endoscopy | Binary | 1,000 | 700/100/200 |
| M2caiSegMSBench<sup>53,54</sup> | Endoscopy | Multi-class (19) | 614 | 245/307/62 |
| MonusacMSBench<sup>55,56</sup> | Pathology | Multi-class (6) | 310 | 188/21/101 |
| MosMedPlusMSBench<sup>57,58</sup> | CT | Binary | 2,729 | 1,910/272/547 |
| NucleiMSBench<sup>59</sup> | Pathology | Binary | 141 | 98/14/29 |
| NusetMSBench<sup>60,61</sup> | Nuclear Cell | Binary | 3,408 | 2,385/340/683 |
| PandentalMSBench<sup>62,63</sup> | X-Ray | Binary | 116 | 81/11/24 |
| PolypGenMSBench<sup>64,65</sup> | Endoscopy | Binary | 1,412 | 984/140/288 |
| Promise12MSBench<sup>66,67</sup> | MRI | Binary | 1,473 | 1,031/147/295 |
| RoboToolMSBench<sup>31</sup> | Endoscopy | Binary | 500 | 350/50/100 |
| TnbcnucleiMSBench<sup>68,69</sup> | Pathology | Binary | 50 | 35/5/10 |
| UltrasoundNerveMSBench<sup>70</sup> | Ultrasound | Binary | 2,323 | 1,651/223/449 |
| USforKidneyMSBench<sup>71,72</sup> | Ultrasound | Binary | 4,586 | 3,210/458/918 |
| UWSkinCancerMSBench<sup>73</sup> | Dermoscopy | Binary | 206 | 143/19/44 |
| WbcMSBench<sup>74,75</sup> | Microscopy | Multi-class (3) | 400 | 280/40/80 |
| YeazMSBench<sup>76,77</sup> | Microscopy | Binary | 707 | 360/96/251 |

**Table 2.**
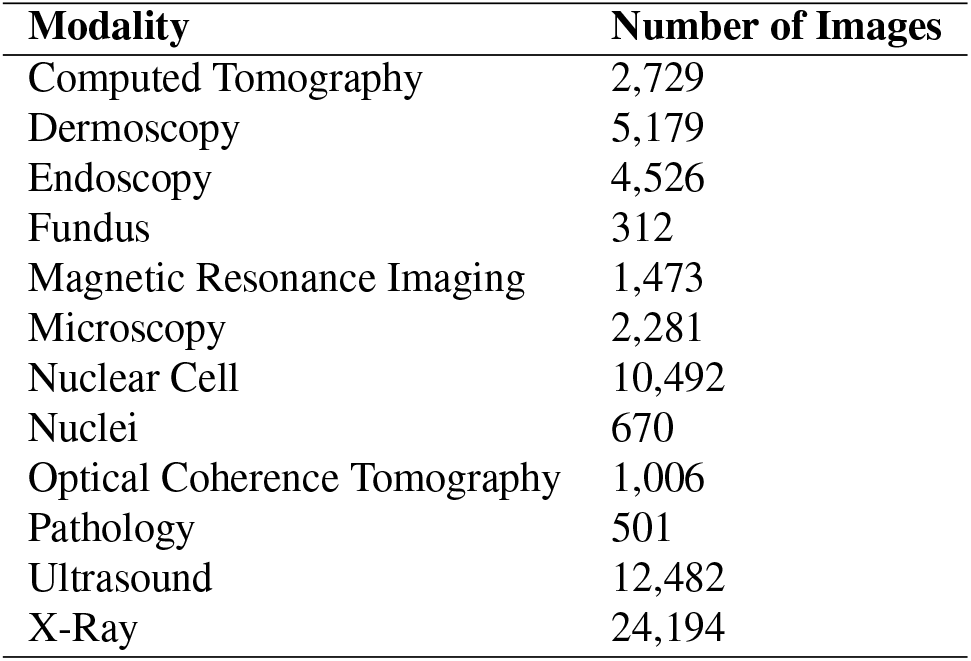
Medical imaging modality and corresponding image counts.

**Table 3.** Overview of datasets and their sub-categories with Train/Validation/Test splits. Each dataset is split into specific sub-categories by authors, and the corresponding number of samples for each sub-category is listed in Train/Val/Test format.

| Dataset Name | Sub-categories | # Train/Val/Test |
| --- | --- | --- |
| BriFiSegMSBench | C1: Target 1 A549;<br>C2: Target 2 A549;<br>C3: HeLa;<br>C4: MCF7;<br>C5: RPE1 | 201/23/48 |
| BusiMSBench | C1: Benign;<br>C2: Malignant | 305/43/89<br>147/21/42 |
| Covid19RadioMSBench | C1: Covid;<br>C2: Lung;<br>C3: Normal;<br>C4: Viral Pneumonia | 2,531/361/724<br>4,208/601/1,203<br>7,134/1,019/2,039<br>941/134/270 |
| IdribMSBench | C1: Microaneurysms;<br>C2: Hemorrhages;<br>C3: Hard Exudates;<br>C4: Optic Disc | 47/6/27 |
| UWSkinCancerMSBench | C1: Melenoma;<br>C2: Not-Melenoma | 83/11/25<br>60/8/19 |
| WbcMSBench | C1: Lymphocyte;<br>C2: Monocyte;<br>C3: Neutrophil;<br>C4: Eosinophil | 146/20/43<br>63/9/43<br>44/6/13<br>23/3/8 |

### Details

#### AbdomenUSMSBench

The AbdomenUSMSBench created from AbdomenUS^11,12^ consists of 926 ultrasound images of the abdominal region, each with a resolution of 449×464 pixels. This dataset is designed for multi-class segmentation tasks and includes eight distinct classes. We have used the official train and test splits, and the train set is split into a training and validation set with a ratio of 9:1. The samples are resized to 1×512×512 pixels, and the labels are mapped to integer values between 0 and (#Classes - 1).

#### Bbbc010MSBench

The Bbbc010MSBench dataset derived from Bbbc010^13,14^, contains 100 microscopy images, each with a resolution of 696×520 pixels. These images are created for binary segmentation tasks and are originally captured for a screen in Fred Ausubel’s Massachusetts General Hospital (MGH) lab. The dataset is split into three parts: train/val/test, in a 7:1:2 ratio. The samples are resized to 1 × 512 × 512 pixels, and the labels are mapped to 0 and 1.

#### Bkai-Igh-MSBench

The Bkai-Igh-MSBench dataset is derived from the BKAI-IGH NeoPolyp dataset^15–17^ and consists of 1,200 endoscopy images, each with a resolution of 1280×995 pixels. It is designed for multi-class segmentation tasks, with three distinct classes. We can not use publicly shared test sets because of a lack of ground truth annotations. The dataset is split into three parts: train/val/test, in a 7:1:2 ratio. The samples are resized to 3×512×512 pixels, and the labels are mapped to integer values between 0 and (#Classes - 1).

#### BriFiSegMSBench

The BriFiSegMSBench, which originates from the BriFiSeg dataset^18,19^, includes 1,360 microscopy images with a resolution of 512×512 pixels. This dataset is intended for binary segmentation tasks and contains two classes. The images are single-channel samples derived from various cell lines, such as A549, HeLa, MCF7, and RPE1. The dataset is divided into training and validation sets with a 9:1 ratio. Additionally, task-specific images and annotations are provided in npz file format (see Table 3). The samples are resized to 1×512×512 pixels, and the labels are mapped to integer values between 0 and 1.

#### BusiMSBench

The BusiMSBench dataset is derived from the Breast Ultrasound Images Dataset^20,21^ and contains 647 ultrasound images with an average resolution of 500×500 pixels. This dataset is designed for binary segmentation tasks, categorizing data into two classes: benign and malignant. It is split into three parts: train/val/test, in a 7:1:2 ratio. Additionally, class-based images (benign and malignant) and annotations are provided in .npz file format (see Table 3). The samples are resized to 1 *×* 512 *×* 512 pixels, and the labels are mapped to integer values between 0 and 1.

#### CellNucleiMSBench

The CellNucleiMSBench comes from the 2018 Data Science Bowl^22,23^ and consists of 670 nuclei images with a resolution of 320×256 pixels. This dataset is specifically designed for binary segmentation tasks. We could not use 65 test images because ground truths are not published officially. Therefore, the source dataset split into three parts: train/val/test, in a 7:1:2 ratio. The samples are resized to 3×512×512 pixels, and the labels are mapped to integer values between 0 and 1.

#### ChaseDB1MSBench

ChaseDB1MSBench is based on the CHASE_DB1 dataset^24^, released in 2012 by Kingston University, London, and St. George’s, University of London, consists of 28 fundus images with a resolution of 999×960 pixels. This dataset is designed for binary segmentation tasks, including two classes. We split the source dataset into three parts: train/val/test, in a 7:1:2 ratio. The samples are resized to 3×512×512 pixels, and the labels are mapped to integer values between 0 and 1.

#### ChuacMSBench

The ChuacMSBench, derived from the CHUAC dataset^25^, includes 28 fundus images with 189×189 pixels. It is designed for binary segmentation tasks. The source dataset is split into three parts: train/val/test, in a 7:1:2 ratio. The samples are resized to 1×512×512 pixels, and the labels are mapped to integer values between 0 and 1.

#### Covid19RadioMSBench

The COVID-19 Radiography Database^26–28^ is the source of the Covid19RadioMSBench dataset, which consists of 21,165 chest X-ray images, each with a resolution of 299×299 pixels. This dataset is designed for binary segmentation tasks. We divide the source dataset into three parts: train/val/test sets with a ratio of 7:1:2. It is developed by a collaborative effort of researchers from Qatar University, the University of Dhaka, and partners from Pakistan and Malaysia, working alongside medical professionals. It includes chest X-ray images for COVID-19 positive cases and Normal and Viral Pneumonia images. The authors have also categorized the images into four groups: COVID, Lung_Opacity, Normal, and Viral Pneumonia. We provide these category-based images and their corresponding annotations in .npz file format (see Table 3). The samples are resized to 1 512 512 pixels, and the labels are mapped to integer values between 0 and 1.

#### CovidQUExMSBench

The CovidQUExMSBench, based on the COVID-QU-Ex Dataset^29,30^, consists of 2,913 chest X-ray images, each with a resolution of 256×256 pixels. This dataset is specifically designed for binary segmentation tasks. We use only infection segmentation samples. The source dataset is split into three parts: train/val/test, in a 7:1:2 ratio. The samples are resized to 1×512×512 pixels, and the labels are mapped to integer values between 0 and 1.

#### MosMedPlusMSBench

The MosMedPlusMSBench, based on the MosMedDataPlus^57,58^ dataset, comprises 2,729 Covid-19 CT images, each sized 512×512 pixels. This dataset is designed for binary segmentation tasks. We split source data into three parts: train/val/test, in a 7:1:2 ratio. The samples are resized to 3×512×512 pixels, and the labels are mapped to integer values between 0 and 1.

#### CystoFluidMSBench

The CystoFluidMSBench is based on Intraretinal Cystoid Fluid dataset^31–33^, comprises 1,006 OCT (Optical Coherence Tomography) images, most of which are sized at 512×512 pixels. This dataset is designed for binary segmentation tasks. The images are carefully chosen by medical experts at Liaquat University of Medical and Health Sciences (LUMHS) Jamshoro, who are trained to identify Cystoid Macular Edema (CME) and its progression, providing a confirmatory diagnosis of CME. The source dataset is split into three parts: train/val/test, in a 7:1:2 ratio. The samples are resized to 3×512×512 pixels, and the labels are mapped to integer values between 0 and 1.

#### Dca1MSBench

The Dca1MSBench is derived from the DCA1 dataset^34,35^ and contains 134 fundus images, each with a resolution of 300×300 pixels. The images are provided by the Cardiology Department of the Mexican Social Security Institute, UMAE T1-León. This dataset is specifically created for binary segmentation tasks. The dataset is split into three parts: train/val/test, in a 7:1:2 ratio. The samples are resized to 1×512×512 pixels, and the labels are mapped to integer values between 0 and 1.

#### DeepbacsMSBench

The DeepbacsMSBench, based on the DeepBacs dataset^36,37^, consists of 34 samples of fundus images, each with a size of 1024×1024 pixels. It is designed for binary segmentation tasks. We use official train/validation/test splits published officially by authors. The samples are resized to 1×512×512 pixels, and the labels are mapped to integer values between 0 and 1.

#### DriveMSBench

The DriveMSBench dataset, based on the DRIVE dataset^38,39^, includes 40 fundus images, each with dimensions of 565×584 pixels. The images are obtained from a diabetic retinopathy screening program in the Netherlands. It is designed for binary segmentation and uses official splits for training, validation, and testing. The samples are resized to 3 *×* 512 *×* 512 pixels, and the labels are mapped to integer values between 0 and 1.

#### DynamicNuclearMSBench

The DynamicNuclearMSBench, created from the DynamicNuclearNet Segmentation dataset40, ^41^, consists of 7084 samples of nuclear cell images, each 128×128 pixels in size. This dataset is utilized for a binary segmentation task. Training, validation, and test splits that are officially published are used. The samples are resized to 1×512×512 pixels, and the labels are mapped to integer values between 0 and 1.

#### FHPsAOPMSBench

The FHPsAOPMSBench dataset is based on a prior dataset^42,43^ and comprises 4,000 ultrasound images, each with a resolution of 256×256 pixels. This dataset is designed for a multi-class segmentation task, including three distinct classes. The source dataset is split into three parts: train/val/test, in a 7:1:2 ratio. The samples are resized to 1 *×* 512 *×* 512 pixels, and the labels are mapped to integer values between 0 and (#Classes - 1).

#### IdribMSBench

The IdribMSBench is based on the Indian Diabetic Retinopathy Image Dataset^44, 45^ and includes 80 high-resolution fundus images (4288×2848 pixels) for a binary segmentation task. We use official train/validation/test splits published officially by authors. The authors have also categorized the labels into four categories: Microaneurysms, hemorrhages, Hard Exudates, and Optic Discs. These category-based labels and annotations are provided in a npz file (see Table 3). The samples are resized to 3×512×512 pixels, and the labels are mapped to integer values between 0 and 1.

#### Isic2016MSBench

The Isic2016MSBench is derived from the ISIC 2016 Challenge^46,47^, which consisted of 1,279 dermoscopy samples of varying sizes designed for binary segmentation tasks. We use official training, validation, and test splits published by authors. The samples are resized to 3×512×512 pixels, and the labels are mapped to integer values between 0 and 1.

#### Isic2018MSBench

The Isic2018MSBench is derived from the ISIC 2018 Challenge^48–50^, which consisted of 3,694 dermoscopy samples of varying sizes designed for binary segmentation tasks. We use official training, validation, and test splits published by authors. The samples are resized to 3×512×512 pixels, and the labels are mapped to integer values between 0 and 1.

#### KvasirMSBench

The KvasirMSBench, derived from the Kvasir-SEG dataset^51,52^, consists of 1,000 endoscopy images with resolutions ranging from 332×487 to 1920×1072 pixels. The dataset includes images of gastrointestinal polyps and their segmentation masks, which have been annotated and verified by an experienced gastroenterologist. It is structured for a binary classification task. The source dataset is divided into three parts: train/val/test, in a 7:1:2 ratio. The samples are resized to 3×512×512 pixels, and the labels are mapped to integer values between 0 and 1.

#### M2caiSegMSBench

M2caiSegMSBench is based on a prior dataset^53,54^ comprising 614 pathology samples and specifically designed for multi-class segmentation tasks, which include 19 distinct classes. The images within this dataset exhibit variable dimensions, and we use official train/validation/test splits. The samples are resized to 3 × 512 ×512 pixels, and the labels are mapped to integer values between 0 and (#Classes - 1).

#### MonusacMSBench

MonusacMSBench is based on the MoNuSAC challenge^55,56^. It consists of 310 samples and is designed for multi-class segmentation with 6 classes. The images in this dataset are H&E stained digitized tissue images from several patients acquired at multiple hospitals using a standard 40x scanner magnification. The annotations are provided by expert pathologists. We use the officially published train/validation/test splits from the challenge. The samples are resized to 3 ×512 ×512 pixels, and the labels are mapped to integer values between 0 and (#Classes - 1).

#### NucleiMSBench

The NucleiMSBench is based on a prior dataset^59^, which consisting of 141 pathology samples each with an image size of 2000×2000 pixels. This source dataset is designed for binary segmentation tasks. The source dataset is split into three parts: train/val/test, in a 7:1:2 ratio. The samples are resized to 3 ×512 ×512 pixels, and the labels are mapped to integer values between 0 and 1.

#### NusetMSBench

The NusetMSBench, derived from the NuSet dataset^60,61^, contains 3,408 pathology samples designed for binary segmentation problems. We split the source dataset into three parts: train/val/test, in a 7:1:2 ratio. The samples are resized to 1 *×* 512 *×* 512 pixels, and the labels are mapped to integer values between 0 and 1.

#### PandentalMSBench

The PandentalMSBench is created from the Panoramic Dental X-rays dataset^62, 63^ and contains 116 X-ray samples of varying sizes. It is specifically intended for binary segmentation tasks. The dataset comprises anonymized and deidentified panoramic dental X-rays of 116 patients taken at Noor Medical Imaging Center in Qom, Iran. The source dataset is divided into three parts: train/val/test, in a 7:1:2 ratio. The samples are resized to 1×512×512 pixels, and the labels are mapped to integer values between 0 and 1.

#### PolypGenMSBench

The PolypGenMSBench is based on a prior endoscopy dataset^64,65^ consisting of 1,412 endoscopy samples, each with an image size of 1920×1080 pixels. It is designed for binary segmentation tasks. It includes colonoscopy video frames captured from a diverse patient population at six different centers in Egypt, France, Italy, Norway, and the United Kingdom. We provide these images and annotations are captured from these centers in a npz file. The source dataset is divided into three parts: train/val/test, in a 7:1:2 ratio. The samples are resized to 3×512×512 pixels, and the labels are mapped to integer values between 0 and 1.

#### Promise12MSBench

The Promise12MSBench, derived on the PROMISE12 dataset^66,67^, contains 1,473 MR samples, each with an image size of 512×512 pixels. It is designed for binary classification. We split the source dataset into three parts: train/val/test, in a 7:1:2 ratio. The samples are resized to 1×512×512 pixels, and the labels are mapped to integer values between 0 and 1.

#### RoboToolMSBench

The RoboToolMSBench, based on the RoboTool dataset^31^, consisting of 500 samples, designed for binary segmentation tasks. The source dataset is divided into three parts: train/val/test, in a 7:1:2 ratio. The samples are resized to 1×512×512 pixels, and the labels are mapped to integer values between 0 and 1.

#### TnbcnucleiMSBench

The TnbcnucleiMSBench is based on a prior dataset^68,69^, consisting of 50 pathology samples, each with an image size of 512×512 pixels. This dataset is based on the merging of two different datasets: the first dataset, generated at the Curie Institute, consists of annotated H&E stained histology images at 40× magnification, and the second dataset, provided by the Indian Institute of Technology Guwahati, also consists of annotated H&E stained histology images captured at 40× magnification. It is designed for binary segmentation tasks. We split the source dataset into three parts: train/val/test, in a 7:1:2 ratio. The samples are resized to 3×512×512 pixels, and the labels are mapped to integer values between 0 and 1.

#### JUltrasoundNerveMSBench

The UltrasoundNerveMSBench, derived from prior dataset^70^, contains 2,323 ultrasound samples, each with an image size of 580×420 pixels and designed for binary segmentation tasks. The primary task in this dataset is to segment a collection of nerves known as the Brachial Plexus (BP) in ultrasound images. Due to the lack of test image annotations, we split the source dataset into three parts: train/val/test, in a 7:1:2 ratio. The samples are resized to 1 *×* 512 *×* 512 pixels, and the labels are mapped to integer values between 0 and 1.

#### USforKidneyMSBench

The USforKidneyMSBench is derived from the CT2USforKidneySeg dataset^71, 72^, comprised of 4,586 ultrasound samples, each with an image size of 256×256 pixels, and designed for binary segmentation tasks. The source dataset is split into three parts: train/val/test, in a 7:1:2 ratio. The samples are resized to 1×512×512 pixels, and the labels are mapped to integer values between 0 and 1.

#### UWSkinCancerMSBench

The UWSkinCancerMSBench is based on the Skin Cancer Detection dataset^73^, consisting of 206 dermoscopy samples, designed for binary classification tasks The dataset includes images extracted from the public databases DermIS and DermQuest, along with manual segmentations of the lesions. We split the source dataset into three parts: train/val/test, in a 7:1:2 ratio. The authors have also categorized the labels into two categories: Melenoma and Not-Melenoma. These category-based labels and annotations are provided in a .npz file (see Table 3). The samples are resized to 3×512×512 pixels, and the labels are mapped to integer values between 0 and 1.

#### WbcMSBench

The WbcMSBench, based on prior datasets^74,75^, is a microscopy imaging dataset consisting of 80 samples, with image sizes of 120×120 and 300×300 pixels. It is designed for multi-class segmentation tasks including 3 classes. The dataset is based on two sources: Dataset 1, obtained from Jiangxi Tecom Science Corporation, China, contains 300 images of white blood cells with a resolution of 120×120 pixels. Dataset 2 consists of 100 color images with a resolution of 300×300 pixels, collected from the CellaVision blog. The authors have grouped the samples into four categories: Lymphocyte, Monocyte, Neutrophil, and Eosinophil, and we provide these category-based images and corresponding labels in npz file format (see Table 3). The source dataset is divided into three parts: train/val/test, in a 7:1:2 ratio. The samples are resized to 3×512×512 pixels, and the labels are mapped to integer values between 0 and (#Classes - 1).

#### YeazMSBench

The YeazMSBench, derived from the YeaZ dataset^76,77^, consists of 707 microscopy images with varying sizes and is designed for binary segmentation tasks. We split the source dataset into three parts: train/val/test, in a 7:1:2 ratio. The samples are resized to 1 × 512 *×* 512 pixels, and the labels are mapped to integer values between 0 and 1.

## Data Records

We have publicly shared each dataset with varying sizes (128, 256, and 512 sized) in MedSegBench at Zenodo (Link). The MedSegBench consists of 35 pre-processed 2D medical image segmentation datasets (some of them extracted 3D slices) from various data modalities and tasks (binary/multi-class). The data storage format published by MedMNISTv2^8^ is followed. We save each dataset in Numpy npz format, named as {dataset}_{size}.npz. Each npz file contains following keys: [“{train,val,test}_images”, “{train,val,test}_label”]. Also, some authors have published class- or category-based images and labels. We have also added this information with the following keys into the npz file and explain them in source code files: [“{train,val,test}_images_ {classno}”, “{train,val,test}_label_{classno}”]. All images and labels are stored in uint8 data type. **{train**,**val**,**test}_images:** Numpy array contains train, validation and test images with *N*×*W*×*H*×*C* shape for RGB datasets, and *N*×*W*×*H* for gray-scale datasets. Here, *N* refers to the number of samples, *W* is the width, *H* is the height, and *C* denotes the number of channels. **{train**,**val**,**test}_label:** It contains train, validation and test labels with *N*×*W*×*H* shape. **{train**,**val**,**test}_images_{classno} and {train**,**val**,**test}_label_{classno}:** These contain class or category-based train, validation, and test images and labels with shapes *N*×*W*×*H*×*C* (for RGB images, and *N*×*W*×*H* for gray-scale images), respectively.

## Technical Validation

### Baseline methods

In this study, we chose the U-Net architecture as the baseline structure for image segmentation tasks. We have selected six encoder/decoder networks to enhance performance and adaptability. These include ResNet18, ResNet50, and DenseNet121, commonly used as benchmarks in segmentation research. Additionally, we have selected EfficientNet and MobileNetv2 because they are lightweight models that offer a more computationally efficient alternative to ResNets and DenseNet. Furthermore, we have added a transformer-based approach using the Mix Vision Transformer, acknowledging the growing interest in transformer models for vision tasks.

The U-Net structure and diverse encoder/decoder networks are implemented using the qubvel-segmentation framework^10^. We have not used pre-trained ImageNet weights; we train each model from scratch on our datasets. We have trained each model with three randomly selected seed values to ensure the robustness of our results. All images are resized to 512×512 pixels, a standardized dimension for the training, validation, and testing phases. Training is conducted over 200 epochs using the Adam Optimizer with a learning rate 1e-3. For binary segmentation tasks, we used dice loss, while categorical cross-entropy loss is used for multi-class tasks. A batch size of 128 is selected throughout the training process. We have not applied weight decay methods or any data augmentation techniques, focusing on the raw performance of the models. The model weights corresponding to the best validation IOU are recorded for each network configuration. Further details regarding the model implementation, training, and evaluation steps are available in our code repository.

### Performance Measures

We have evaluated each model on 35 different datasets using four performance measures: Precision (PREC), Recall (REC), F1-score (F1), and Intersection over Union (IOU). Precision measures the accuracy of positive predictions, highlighting its ability to avoid false positives, while Recall evaluates the model’s capacity to identify all relevant positive instances, minimizing false negatives. The F1-Score, as the harmonic mean of Precision and Recall, provides a balanced view, which is especially useful when there is an unbalanced class distribution. IoU, primarily used in image segmentation and object detection, evaluates the overlap between predicted and actual regions, ensuring accurate localization and identification of objects. We have individually reported PREC, REC, F1, and IOU scores for each dataset and averaged the results.

## Results

The average PREC and REC results obtained from three different run are showed in Table 4 and average F1 and IOU scores are reported in Table 5 for each individual datasets. Also, the average results for each baseline methods are shown in Table 5,

**Table 4.**
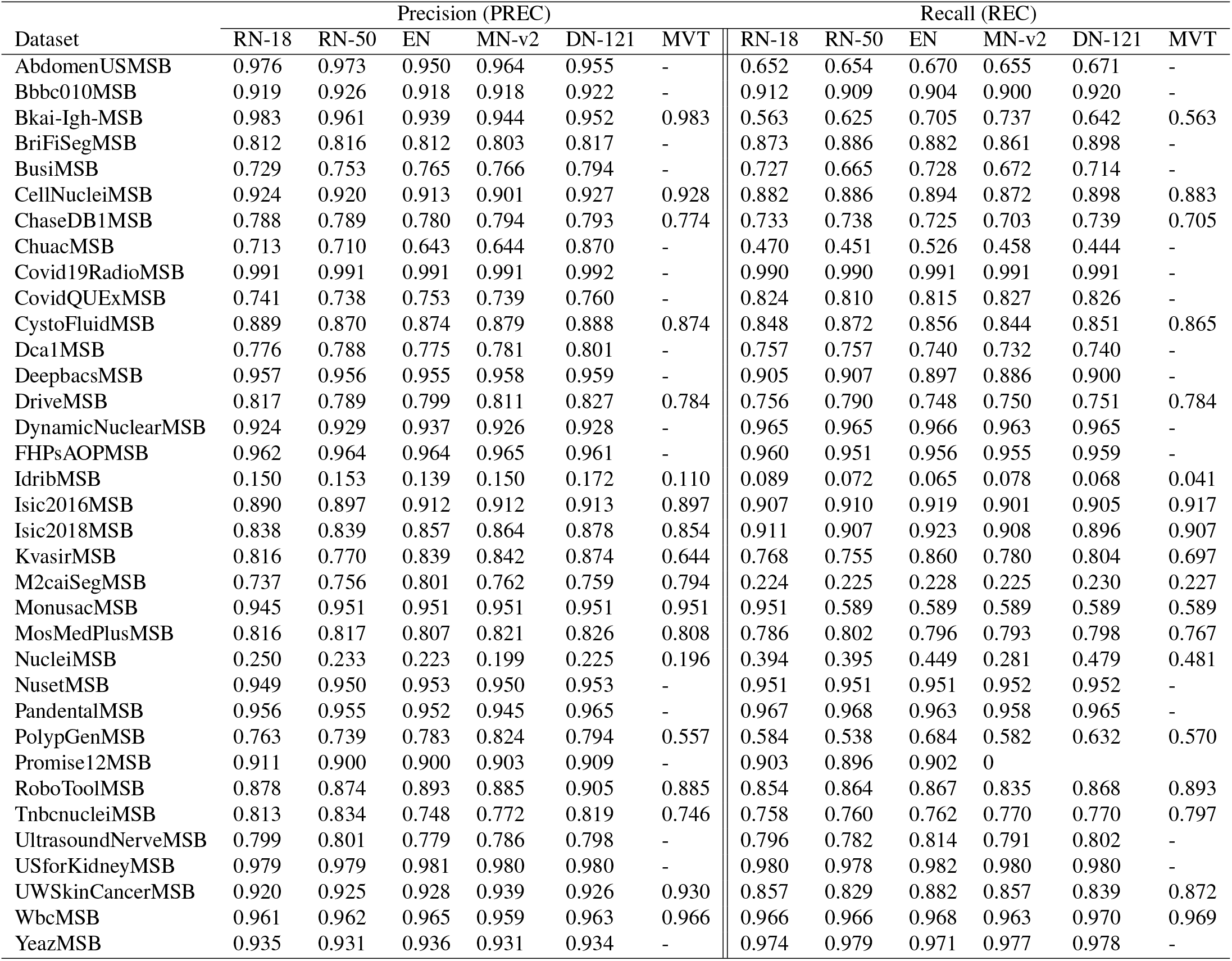
The average precision and recall results for six different encoder networks. RN-18: ResNet-18; RN-50: ResNet-50; EN: Efficient-Net; MN-v2: Mobile-Net-v2; DN-121: DenseNet-121; MVT: Mix Vision Transformer

**Table 5.** The average F1-score and IOU results for six different encoder networks. RN-18: ResNet-18; RN-50: ResNet-50; EN: Efficient-Net; MN-v2: Mobile-Net-v2; DN-121: DenseNet-121; MVT: Mix Vision Transformer

| Dataset | F1-Score (F1) |  |  |  |  |  | Intersection over Union (IOU) |  |  |  |  |  |
| --- | --- | --- | --- | --- | --- | --- | --- | --- | --- | --- | --- | --- |
|  | RN-18 | RN-50 | EN | MN-v2 | DN-121 | MVT | RN-18 | RN-50 | EN | MN-v2 | DN-121 | MVT |
| AbdomenUSMSB | 0.642 | 0.640 | 0.640 | 0.635 | 0.643 | - | 0.632 | 0.630 | 0.628 | 0.624 | 0.632 | - |
| Bbbc010MSB | 0.915 | 0.917 | 0.910 | 0.908 | 0.920 | - | 0.844 | 0.848 | 0.837 | 0.833 | 0.854 | - |
| Bkai-Igh-MSB | 0.554 | 0.617 | 0.692 | 0.733 | 0.630 | 0.554 | 0.546 | 0.604 | 0.676 | 0.713 | 0.615 | 0.546 |
| BriFiSegMSB | 0.826 | 0.834 | 0.831 | 0.816 | 0.840 | - | 0.717 | 0.728 | 0.724 | 0.702 | 0.738 | - |
| BusiMSB | 0.674 | 0.632 | 0.711 | 0.655 | 0.695 | - | 0.578 | 0.547 | 0.624 | 0.565 | 0.615 | - |
| CellNucleiMSB | 0.889 | 0.892 | 0.894 | 0.880 | 0.907 | 0.891 | 0.822 | 0.827 | 0.830 | 0.815 | 0.838 | 0.826 |
| ChaseDB1MSB | 0.758 | 0.761 | 0.750 | 0.744 | 0.764 | 0.735 | 0.611 | 0.615 | 0.601 | 0.594 | 0.618 | 0.582 |
| ChuacMSB | 0.487 | 0.451 | 0.499 | 0.462 | 0.522 | - | 0.357 | 0.334 | 0.369 | 0.340 | 0.400 | - |
| Covid19RadioMSB | 0.991 | 0.990 | 0.991 | 0.991 | 0.992 | - | 0.982 | 0.981 | 0.983 | 0.982 | 0.983 | - |
| CovidQUExMSB | 0.740 | 0.734 | 0.744 | 0.742 | 0.756 | - | 0.627 | 0.620 | 0.633 | 0.631 | 0.647 | - |
| CystoFluidMSB | 0.852 | 0.857 | 0.849 | 0.842 | 0.853 | 0.855 | 0.759 | 0.765 | 0.754 | 0.747 | 0.761 | 0.763 |
| Dca1MSB | 0.762 | 0.767 | 0.753 | 0.751 | 0.765 | - | 0.618 | 0.625 | 0.606 | 0.604 | 0.623 | - |
| DeepbacsMSB | 0.930 | 0.931 | 0.925 | 0.921 | 0.929 | - | 0.869 | 0.870 | 0.860 | 0.853 | 0.867 | - |
| DriveMSB | 0.782 | 0.786 | 0.770 | 0.775 | 0.782 | 0.781 | 0.643 | 0.648 | 0.626 | 0.634 | 0.643 | 0.641 |
| DynamicNuclearMSB | 0.941 | 0.942 | 0.948 | 0.940 | 0.942 | - | 0.895 | 0.897 | 0.906 | 0.893 | 0.897 | - |
| FHPsAOPMSB | 0.961 | 0.957 | 0.959 | 0.959 | 0.960 | - | 0.929 | 0.923 | 0.927 | 0.927 | 0.928 | - |
| IdribMSB | 0.100 | 0.090 | 0.078 | 0.092 | 0.089 | 0.053 | 0.061 | 0.054 | 0.046 | 0.056 | 0.054 | 0.030 |
| Isic2016MSB | 0.878 | 0.887 | 0.903 | 0.891 | 0.893 | 0.891 | 0.803 | 0.814 | 0.836 | 0.820 | 0.825 | 0.822 |
| Isic2018MSB | 0.849 | 0.849 | 0.868 | 0.865 | 0.861 | 0.853 | 0.761 | 0.762 | 0.790 | 0.783 | 0.785 | 0.773 |
| KvasirMSB | 0.739 | 0.698 | 0.812 | 0.754 | 0.794 | 0.569 | 0.645 | 0.596 | 0.733 | 0.668 | 0.718 | 0.457 |
| M2caiSegMSB | 0.214 | 0.215 | 0.218 | 0.216 | 0.223 | 0.217 | 0.190 | 0.191 | 0.196 | 0.192 | 0.200 | 0.194 |
| MonusacMSB | 0.557 | 0.559 | 0.559 | 0.559 | 0.559 | 0.538 | 0.540 | 0.540 | 0.540 | 0.540 | 0.540 | 0.540 |
| MosMedPlusMSB | 0.780 | 0.790 | 0.781 | 0.785 | 0.791 | 0.761 | 0.674 | 0.682 | 0.674 | 0.679 | 0.686 | 0.650 |
| NucleiMSB | 0.282 | 0.274 | 0.278 | 0.205 | 0.275 | 0.253 | 0.169 | 0.164 | 0.167 | 0.119 | 0.166 | 0.150 |
| NusetMSB | 0.949 | 0.949 | 0.951 | 0.950 | 0.951 | - | 0.906 | 0.906 | 0.909 | 0.907 | 0.910 | - |
| PandentalMSB | 0.961 | 0.961 | 0.957 | 0.950 | 0.965 | - | 0.926 | 0.926 | 0.919 | 0.907 | 0.932 | - |
| PolypGenMSB | 0.573 | 0.541 | 0.666 | 0.588 | 0.621 | 0.477 | 0.495 | 0.457 | 0.587 | 0.512 | 0.545 | 0.382 |
| Promise12MSB | 0.895 | 0.888 | 0.892 | 0.896 | 0.900 | - | 0.828 | 0.817 | 0.821 | 0.827 | 0.832 | - |
| RoboToolMSB | 0.856 | 0.859 | 0.874 | 0.847 | 0.879 | 0.882 | 0.765 | 0.769 | 0.788 | 0.753 | 0.798 | 0.798 |
| TnbnucleiMSB | 0.779 | 0.785 | 0.738 | 0.762 | 0.788 | 0.759 | 0.641 | 0.652 | 0.596 | 0.621 | 0.654 | 0.618 |
| UltrasoundNerveMSB | 0.782 | 0.776 | 0.787 | 0.772 | 0.786 | - | 0.671 | 0.664 | 0.675 | 0.660 | 0.676 | - |
| USforKidneyMSB | 0.979 | 0.978 | 0.981 | 0.980 | 0.980 | - | 0.960 | 0.958 | 0.963 | 0.961 | 0.960 | - |
| UWSkinCancerMSB | 0.864 | 0.846 | 0.890 | 0.879 | 0.856 | 0.881 | 0.795 | 0.766 | 0.818 | 0.803 | 0.779 | 0.813 |
| WbcMSB | 0.962 | 0.963 | 0.966 | 0.959 | 0.966 | 0.967 | 0.930 | 0.931 | 0.937 | 0.926 | 0.936 | 0.938 |
| YeazMSB | 0.953 | 0.953 | 0.952 | 0.952 | 0.954 | - | 0.912 | 0.912 | 0.909 | 0.910 | 0.914 | - |

Table 4 presents a comprehensive overview of the average precision and recall results for six different encoder networks across various datasets. These networks include ResNet-18 (RN-18), ResNet-50 (RN-50), Efficient-Net (EN), Mobile-Net-v2 (MN-v2), DenseNet-121 (DN-121), and Mix Vision Transformer (MVT). The results are divided into two main categories: precision and recall. In terms of precision, DenseNet-121 consistently demonstrated strong performance across numerous datasets. For example, it achieved the highest precision scores in datasets such as BusiMSB (0.794), ChuahMSB(0.870) and Dca1MSB (0.801). Similarly, Efficient-Net also demonstrated strong precision, particularly in datasets like Isic2016MSB and Isic2018MSB, where it scored 0.912 and 0.857, respectively. Although the Mix Vision Transformer is not evaluated on all datasets because it only accepts at least three channel images as input, it performed competitively where applicable, achieving high precision in datasets like Bkai-Igh-MSB (0.983). Regarding Recall, DenseNet-121 has emerged as a top performer, achieving the highest recall in datasets such as Bbbbc010MSB (0.920) and WbcMSB (0.970). Efficient-Net also performed well in recall metric, particularly in datasets like DynamicNuclearMSB (0.966) and USforKidneyMSB (0.982). The results indicate that DenseNet-121 and Efficient-Net are particularly robust across precision and recall metrics, suggesting their effectiveness in various applications. Overall, the analysis highlights DenseNet-121’s consistently high performance across multiple datasets, making it a reliable choice for tasks requiring high precision and recall. Efficient-Net also stands out, especially in recall, indicating its potential for applications where recall is critical.

Table 5 provides a comprehensive evaluation of six encoder networks across various datasets, using F1-score and Intersection over Union (IOU) as performance metrics. DenseNet-121 consistently performs well, frequently achieving the top F1 and IOU metrics scores across numerous datasets. For example, in the Bbbc010MSB and CellNucleiMSB datasets, DenseNet-121 records the highest F1-scores of 0.920 and 0.907, respectively, and similarly high IOU scores, indicating its robustness in handling diverse data types. Efficient-Net also shows significant performance, particularly in datasets like Isic2016MSB and USforKidneyMSB, where it achieves the highest F1-scores of 0.903 and 0.981, respectively. This indicates that Efficient-Net is particularly effective in scenarios requiring high precision and recall, as showed in its F1-scores. ResNet-50 performs best with an F1-score of 0.931 and an IOU of 0.870 for the DeepbacsMSB. Additionally, it has also achieved the highest F1-score of 0.786 and an IOU of 0.648 in the DriveMSB dataset. For the FHPsAOPMSB dataset, ResNet-18 has achieved the highest F1-score of 0.961 and an IOU of 0.929. While Mix Vision Transformer does not frequently perform as well as DenseNet-121, it shows competitive performance in specific datasets such as UWSkinCancerMSB, achieving the second-highest F1 Score of 0.881. This indicates its potential in specialized applications, particularly in medical imaging contexts. Overall, DenseNet-121 is the most robust and effective network, frequently outperforming other networks in achieving high F1-scores and IOU values.

Table 6 shows the average performance metrics for six different encoder networks. Efficient-Net (EN) and DenseNet-121 (DN-121) demonstrate the highest F1 scores, both achieving a value of 0.772. This indicates that these models have a balanced performance in terms of precision and recall. DenseNet-121 also achieves the highest precision at 0.848, suggesting it is effective at minimizing false positives. On the other hand, Efficient-Net leads in recall with a score of 0.788, indicating its strength in capturing true positives. Additionally, DenseNet-121 achieves the highest IOU with 0.702, closely followed by Efficient-Net with 0.700 This suggests that these two models provide the most accurate predictions. Overall, DenseNet-121 and Efficient-Net achieve similar high-performance metrics, with both models performing well in F1 score, precision, recall, and IOU. However, DenseNet-121’s complex architecture causes higher computational demands, whereas Efficient-Net provides a more efficient design, making it suitable for resource-constrained applications.

**Table 6.** The average results for six different encoder networks. RN-18: ResNet-18; RN-50: ResNet-50; EN: Efficient-Net; MN-v2: Mobile-Net-v2; DN-121: DenseNet-121; MVT: Mix Vision Transformer.

| Methods | F1 | PREC | REC | IOU |
| --- | --- | --- | --- | --- |
| RN-18 | 0.762 | 0.834 | 0.774 | 0.689 |
| RN-50 | 0.759 | 0.833 | 0.772 | 0.686 |
| EN | **0.772** | 0.832 | **0.788** | 0.700 |
| MN-v2 | 0.762 | 0.834 | 0.769 | 0.689 |
| DN-121 | **0.772** | **0.848** | 0.781 | **0.702** |
| MVT | 0.663 | 0.760 | 0.696 | 0.585 |

### Usage Notes

The MedSegBench datasets are freely available at Zenodo We kindly request that users of the MedSegBench dataset cite this paper, along with the relevant source dataset files, in their publications. This dataset is created in order to fairly compare different models over various segmentation models from different data modalities and to create universal models. It is not suitable for clinical or medical use.

## Data Availability

All data produced are available online at Zenodo

https://zenodo.org/records/13359660

## Code availability

The Python data API, source code files and evaluation scripts for binary and multi-class segmentation tasks can be found at https://github.com/zekikus/MedSegBench.

## Acknowledgements

We would like to express our gratitude for the authors of the MedMNIST^8^, which served as the baseline for our study, and for the shared source code that we referenced to develop our own code.

## Author contributions statement

M.A. conducted data collection, cleaning and pre-processing steps. Z.K. performed the evaluation tests for binary and multi-class task for each network and datasets. All authors wrote and reviewed the manuscript.

## References

1. Han, K. et al. Deep semi-supervised learning for medical image segmentation: A review. Expert. Syst. with Appl. 245, 123052, 10.1016/j.eswa.2023.123052 (2024).

2. Ma, J. et al. Segment anything in medical images. Nat. Commun. 15, 10.1038/s41467-024-44824-z (2024).

3. Carriero, A., Groenhoff, L., Vologina, E., Basile, P. & Albera, M. Deep learning in breast cancer imaging: State of the art and recent advancements in early 2024. Diagnostics 14, 848, 10.3390/diagnostics14080848 (2024).

4. Drelie Gelasca, E., Obara, B., Fedorov, D., Kvilekval, K. & Manjunath, B. A biosegmentation benchmark for evaluation of bioimage analysis methods. BMC Bioinforma. 10, 10.1186/1471-2105-10-368 (2009).

5. Rebuffi, S.-A., Bilen, H. & Vedaldi, A. Learning multiple visual domains with residual adapters. In Guyon, I.et al. (eds.) Advances in Neural Information Processing Systems, vol. 30 (Curran Associates, Inc., 2017).

6. Simpson, A. L. et al. A large annotated medical image dataset for the development and evaluation of segmentation algorithms. CoRR abs/1902.09063 (2019). 1902.09063.

7. Yang, J., Shi, R. & Ni, B. Medmnist classification decathlon: A lightweight automl benchmark for medical image analysis. In 2021 IEEE 18th International Symposium on Biomedical Imaging (ISBI), 10.1109/isbi48211.2021.9434062 (IEEE, 2021).

8. Yang, J. et al. Medmnist v2 - a large-scale lightweight benchmark for 2d and 3d biomedical image classification. Sci. Data 10, 10.1038/s41597-022-01721-8 (2023).

9. Ronneberger, O., Fischer, P. & Brox, T. U-Net: Convolutional Networks for Biomedical Image Segmentation, 234–241 (Springer International Publishing, 2015).

10. Iakubovskii, P. Segmentation models pytorch. https://github.com/qubvel/segmentation_models.pytorch (2019).

11. Vitale, S., Orlando, J. I., Iarussi, E. & Larrabide, I. Improving realism in patient-specific abdominal ultrasound simulation using cyclegans. Int. J. Comput. Assist. Radiol. Surg. 15, 183–192, 10.1007/s11548-019-02046-5 (2019).

12. Orlando, J. I. Us simulation & segmentation (2020).

13. Ljosa, V., Sokolnicki, K. L. & Carpenter, A. E. Annotated high-throughput microscopy image sets for validation. Nat. Methods 9, 637–637, 10.1038/nmeth.2083 (2012).

14. Broad Bioimage Benchmark Collection — bbbc.broadinstitute.org. https://bbbc.broadinstitute.org/BBBC010. [Accessed 06-08-2024].

15. Ngoc Lan, P. et al. NeoUNet: Towards Accurate Colon Polyp Segmentation and Neoplasm Detection, 15–28 (Springer International Publishing, 2021).

16. An, N. S. et al. Blazeneo: Blazing fast polyp segmentation and neoplasm detection. IEEE Access 10, 43669–43684, 10.1109/access.2022.3168693 (2022).

17. Duc, N. T., Oanh, N. T., Thuy, N. T., Triet, T. M. & Dinh, V. S. Colonformer: An efficient transformer based method for colon polyp segmentation. IEEE Access 10, 80575–80586, 10.1109/access.2022.3195241 (2022).

18. Mathieu, G. M. L. A., & Bachir, E. D. Brifiseg: a deep learning-based method for semantic and instance segmentation of nuclei in brightfield images, 10.48550/ARXIV.2211.03072 (2022).

19. Gendarme, M. & Debs, B. E. Brifiseg datasets, 10.5281/ZENODO.7195636 (2022).

20. Al-Dhabyani, W., Gomaa, M., Khaled, H. & Fahmy, A. Dataset of breast ultrasound images. Data Brief 28, 104863, 10.1016/j.dib.2019.104863 (2020).

21. Breast Ultrasound Images Dataset — kaggle.com. https://www.kaggle.com/datasets/aryashah2k/breast-ultrasound-images-dataset. [Accessed 06-08-2024].

22. Caicedo, J. C. et al. Nucleus segmentation across imaging experiments: the 2018 data science bowl. Nat. Methods 16, 1247–1253, 10.1038/s41592-019-0612-7 (2019).

23. 2018 Data Science Bowl — kaggle.com. https://www.kaggle.com/competitions/data-science-bowl-2018/data. [Accessed 06-08-2024].

24. Carballal, A. et al. Automatic multiscale vascular image segmentation algorithm for coronary angiography. Biomed. Signal Process. Control. 46, 1–9, 10.1016/j.bspc.2018.06.007 (2018).

25. Angiographics — figshare.com. https://figshare.com/s/4d24cf3d14bc901a94bf. [Accessed 06-08-2024].

26. Chowdhury, M. E. H. et al. Can ai help in screening viral and covid-19 pneumonia? IEEE Access 8, 132665–132676, 10.1109/access.2020.3010287 (2020).

27. Rahman, T. et al. Exploring the effect of image enhancement techniques on covid-19 detection using chest x-ray images. Comput. Biol. Medicine 132, 104319, 10.1016/j.compbiomed.2021.104319 (2021).

28. COVID-19 Radiography Database — kaggle.com. https://www.kaggle.com/datasets/tawsifurrahman/ covid19-radiography-database. [Accessed 06-08-2024].

29. Tahir, A. M. et al. Covid-19 infection localization and severity grading from chest x-ray images. Comput. Biol. Medicine 139, 105002, 10.1016/j.compbiomed.2021.105002 (2021).

30. Anas M. Tahir et al. Covid-qu-ex dataset, 10.34740/KAGGLE/DSV/3122958 (2022).

31. Garcia-Peraza-Herrera, L. C. et al. Image compositing for segmentation of surgical tools without manual annotations. IEEE Transactions on Med. Imaging 40, 1450–1460, 10.1109/tmi.2021.3057884 (2021).

32. Zeeshan Ahmed, Munawar Ahmed, Attiya Baqai & Fahim Aziz Umrani. Intraretinal cystoid fluid, 10.34740/KAGGLE/DS/2277068 (2022).

33. Ahmed, Z. et al. Deep learning based automated detection of intraretinal cystoid fluid. Int. J. Imaging Syst. Technol. 32, 902–917, 10.1002/ima.22662 (2021).

34. Cervantes-Sanchez, F., Cruz-Aceves, I., Hernandez-Aguirre, A., Hernandez-Gonzalez, M. A. & Solorio-Meza, S. E. Automatic segmentation of coronary arteries in x-ray angiograms using multiscale analysis and artificial neural networks. Appl. Sci. 9, 5507, 10.3390/app9245507 (2019).

35. Ivan Cruz Aceves CIMAT — personal.cimat.mx. http://personal.cimat.mx:8181/~ivan.cruz/DB_Angiograms.html. [Accessed 06-08-2024].

36. Spahn, C. et al. Deepbacs for multi-task bacterial image analysis using open-source deep learning approaches. Commun. Biol. 5, 10.1038/s42003-022-03634-z (2022).

37. Spahn, C. & Heilemann, M. Deepbacs – escherichia coli bright field segmentation dataset, 10.5281/ZENODO.5550934 (2021).

38. Staal, J., Abramoff, M., Niemeijer, M., Viergever, M. & van Ginneken, B. Ridge-based vessel segmentation in color images of the retina. IEEE Transactions on Med. Imaging 23, 501–509, 10.1109/tmi.2004.825627 (2004).

39. DRIVE - Grand Challenge — drive.grand-challenge.org. https://drive.grand-challenge.org/. [Accessed 06-08-2024].

40. Van Valen, D. A. et al. Deep learning automates the quantitative analysis of individual cells in live-cell imaging experiments. PLOS Comput. Biol. 12, e1005177, 10.1371/journal.pcbi.1005177 (2016).

41. DeepCell Datasets — datasets.deepcell.org. https://datasets.deepcell.org/data. [Accessed 06-08-2024].

42. Lu, Y. et al. The jnu-ifm dataset for segmenting pubic symphysis-fetal head. Data Brief 41, 107904, 10.1016/j.dib.2022.107904 (2022).

43. Jieyun, B. & ZhanHong, O. Pubic symphysis-fetal head segmentation and angle of progression, 10.5281/ZENODO.7851338 (2024).

44. Porwal, P. et al. Indian diabetic retinopathy image dataset (idrid): A database for diabetic retinopathy screening research. Data 3, 25, 10.3390/data3030025 (2018).

45. Prasanna Porwal, S. P. Indian diabetic retinopathy image dataset (idrid), 10.21227/H25W98 (2018).

46. Codella, N. C. F. et al. Skin lesion analysis toward melanoma detection: A challenge at the 2017 international symposium on biomedical imaging (isbi), hosted by the international skin imaging collaboration (isic). In 2018 IEEE 15th International Symposium on Biomedical Imaging (ISBI 2018), 10.1109/isbi.2018.8363547 (IEEE, 2018).

47. ISIC Challenge — challenge.isic-archive.com. https://challenge.isic-archive.com/data/#2016. [Accessed 07-08-2024].

48. Tschandl, P., Rosendahl, C. & Kittler, H. The ham10000 dataset, a large collection of multi-source dermatoscopic images of common pigmented skin lesions. Sci. Data 5, 10.1038/sdata.2018.161 (2018).

49. Codella, N. et al. Skin lesion analysis toward melanoma detection 2018: A challenge hosted by the international skin imaging collaboration (isic), 10.48550/ARXIV.1902.03368 (2019).

50. ISIC Challenge — challenge.isic-archive.com. https://challenge.isic-archive.com/data/#2018. [Accessed 07-08-2024].

51. Jha, D. et al. Kvasir-SEG: A Segmented Polyp Dataset, 451–462 (Springer International Publishing, 2019).

52. Simula Datasets - Kvasir SEG — datasets.simula.no. https://datasets.simula.no/kvasir-seg/. [Accessed 06-08-2024].

53. Maqbool, S., Riaz, A., Sajid, H. & Hasan, O. m2caiseg: Semantic segmentation of laparoscopic images using convolutional neural networks, 10.48550/ARXIV.2008.10134 (2020).

54. m2caiSeg — kaggle.com. https://www.kaggle.com/datasets/salmanmaq/m2caiseg. [Accessed 07-08-2024].

55. Verma, R. et al. Monusac2020: A multi-organ nuclei segmentation and classification challenge. IEEE Transactions on Med. Imaging 40, 3413–3423, 10.1109/tmi.2021.3085712 (2021).

56. MoNuSAC 2020 - Grand Challenge — monusac-2020.grand-challenge.org. https://monusac-2020.grand-challenge.org/Data/. [Accessed 07-08-2024].

57. Morozov, S. P. et al. Mosmeddata: Chest ct scans with covid-19 related findings dataset, 10.48550/ARXIV.2005.06465 (2020).

58. COVID-19 CT scan lesion segmentation dataset — kaggle.com. https://www.kaggle.com/datasets/maedemaftouni/ covid19-ct-scan-lesion-segmentation-dataset. [Accessed 06-08-2024].

59. Janowczyk, A. & Madabhushi, A. Deep learning for digital pathology image analysis: A comprehensive tutorial with selected use cases. J. Pathol. Informatics 7, 29, 10.4103/2153-3539.186902 (2016).

60. Yang, L. et al. Nuset: A deep learning tool for reliably separating and analyzing crowded cells. PLOS Comput. Biol. 16, e1008193, 10.1371/journal.pcbi.1008193 (2020).

61. Linfeng Yang. Nuset training dataset/model weights from (nuset: A deep learning tool for reliably separating and analyzing crowded cells), 10.5281/ZENODO.3996369 (2020).

62. Abdi, A. H., Kasaei, S. & Mehdizadeh, M. Automatic segmentation of mandible in panoramic x-ray. J. Med. Imaging 2, 044003, 10.1117/1.jmi.2.4.044003 (2015).

63. Abdi, A. Panoramic dental x-rays with segmented mandibles, 10.17632/HXT48YK462.1 (2017).

64. Ali, S. et al. Assessing generalisability of deep learning-based polyp detection and segmentation methods through a computer vision challenge, 10.48550/ARXIV.2202.12031 (2022).

65. Ali, S. et al. Deep learning for detection and segmentation of artefact and disease instances in gastrointestinal endoscopy. Med. Image Analysis 70, 102002, 10.1016/j.media.2021.102002 (2021).

66. Litjens, G. et al. Evaluation of prostate segmentation algorithms for mri: The promise12 challenge. Med. Image Analysis 18, 359–373, 10.1016/j.media.2013.12.002 (2014).

67. Litjens, G. et al. Promise12: Data from the miccai grand challenge: Prostate mr image segmentation 2012, 10.5281/ZENODO.8014040 (2023).

68. Jack, N. P., Thomas, W., Laé Marick & Reyal Fabien. Segmentation of nuclei in histopathology images by deep regression of the distance map, 10.5281/ZENODO.1175282 (2018).

69. Naylor, P., Laé, M., Reyal, F. & Walter, T. Segmentation of nuclei in histopathology images by deep regression of the distance map. IEEE Transactions on Med. Imaging 38, 448–459, 10.1109/tmi.2018.2865709 (2019).

70. Ultrasound Nerve Segmentation — kaggle.com. https://www.kaggle.com/competitions/ultrasound-nerve-segmentation. [Accessed 07-08-2024].

71. Song, Y. et al. Ct2us: Cross-modal transfer learning for kidney segmentation in ultrasound images with synthesized data. Ultrasonics 122, 106706, 10.1016/j.ultras.2022.106706 (2022).

72. CT2USforKidneySeg — kaggle.com. https://www.kaggle.com/datasets/siatsyx/ct2usforkidneyseg/data. [Accessed 07-08-2024].

73. Skin Cancer Detection | Vision and Image Processing Lab — uwaterloo.ca. https://uwaterloo.ca/ vision-image-processing-lab/research-demos/skin-cancer-detection. [Accessed 07-08-2024].

74. Zheng, X., Wang, Y., Wang, G. & Liu, J. Fast and robust segmentation of white blood cell images by self-supervised learning. Micron 107, 55–71, 10.1016/j.micron.2018.01.010 (2018).

75. Acevedo, A., Alférez, S., Merino, A., Puigví, L. & Rodellar, J. Recognition of peripheral blood cell images using convolutional neural networks. Comput. Methods Programs Biomed. 180, 105020, 10.1016/j.cmpb.2019.105020 (2019).

76. Dietler, N. et al. A convolutional neural network segments yeast microscopy images with high accuracy. Nat. Commun. 11, 10.1038/s41467-020-19557-4 (2020).

77. Data and Software — epfl.ch. https://www.epfl.ch/labs/lpbs/data-and-software/. [Accessed 07-08-2024].

